# Severe Outcomes in Pediatric Gastrointestinal Infections Presenting to Emergency Departments: A National Surveillance Study

**DOI:** 10.1101/2025.11.24.25340803

**Authors:** Aniruth Ananthanarayanan, Benjamin Hu, Tarun Achar

**Affiliations:** Texas Academy of Mathematics and Science, University of North Texas, Denton, TX, United States of America

## Abstract

**Introduction:** Pediatric gastrointestinal (GI) infections are a major cause of emergency department (ED) visits, yet the epidemiology of severe cases requiring hospitalization or intensive care remains poorly characterized. This study assessed the incidence, predictors, and temporal patterns of severe pediatric GI infections using national surveillance data.

**Methods:** We analyzed 7,326,429 records from the National Electronic Injury Surveillance System (NEISS) for patients ≤ 18 years with narrative descriptions (n=2,876,152). GI infections were identified using keywords including vomiting, diarrhea, nausea, food poisoning, and pathogens such as Salmonella, E. coli, norovirus, Giardia, Campylobacter, and Shigella. Severe cases were defined by hospitalization, ICU admission, intravenous fluid administration, or hospital transfer. Weighted incidence rates were calculated per 100,000 ED visits. Multivariable logistic regression adjusting for year, age, sex, and month evaluated associations with severe outcomes. Seasonal and yearly trends were also examined.

**Results:** Among pediatric ED visits, 438,354 weighted cases (0.50%) involved GI infections. Severe outcomes occurred in 0.82% of GI infection cases versus 0.21% in non-GI visits, equating to 8,179 versus 2,068 severe cases per 100,000 ED visits. GI infection was strongly associated with severe outcomes (adjusted OR=3.95, 95% CI: 3.82–4.09, p<0.001). Younger age (OR=0.99 per year, p<0.001) and male sex were also significant predictors. Seasonal peaks were observed in March, with troughs in May and December.

**Conclusions:** Pediatric GI infections carry nearly four-fold increased odds of severe outcomes, highlighting a substantial burden. Seasonal variability underscores the need for preventive measures, food safety initiatives, and targeted public health interventions to reduce severe pediatric GI infections.

## INTRODUCTION

Although gastrointestinal infections represent one of the most common reasons for pediatric emergency department visits in the United States,^1,2^ comprehensive data on severe outcomes requiring hospitalization remain limited. Most prior studies examining pediatric GI infection burden have been small, single-institution investigations^3,4^ or focused on specific pathogens detected through laboratory testing. Additionally, many studies predated recent shifts in pathogen epidemiology following rotavirus vaccine introduction and changes in foodborne illness patterns, thus excluding important contemporary trends.^5-7^

Several national surveillance systems track GI infection incidence,^3,8^ but few have systematically characterized severe outcome patterns across the full spectrum of pediatric presentations to emergency departments. Whether findings from pathogen-specific studies apply to the broader population of children presenting with GI symptoms remains unknown. To our knowledge, no study has comprehensively examined severe outcome incidence comparing screen-detected versus clinically symptomatic pediatric GI infections. With increasing emergency department utilization and the corresponding increase in the number of children diagnosed with GI infections,^9,10^ a better understanding of severe outcome patterns and predictors has become paramount to optimizing clinical management and improving outcomes.

The objective of this study was to examine the incidence, timing, and severe outcomes of pediatric gastrointestinal infections using data from the National Electronic Injury Surveillance System.

## METHODS

### Data source and study population

We analyzed records from the U.S. Consumer Product Safety Commission’s National Electronic Injury Surveillance System (NEISS). The analysis used all NEISS files from 2005-2024 and included encounters from all years present in those files. A total of 7,326,429 NEISS records were loaded for processing. We restricted the analytic sample to records for patients aged ≤18 years and to encounters with a non-empty clinical narrative field; after these restrictions the analytic sample comprised 2,876,152 unique ED encounter records.

### Case definition and text-based classification

Because NEISS does not include routine laboratory-confirmation fields for infectious disease, gastrointestinal (GI) infection case status was inferred by text search of the clinical narrative. An a priori keyword list was used to identify probable GI infections; the keyword list used in the script was: vomiting, diarrhea, nausea, food poisoning, salmonella, E.coli, norovirus, giardia, campylobacter, shigella (case-insensitive, whole-text regular expression match). Records whose narrative matched any of these terms were labeled as being related to GI infections.

### Definition of severe outcome

A severe ED outcome was defined by evidence of hospital-level care documented in the NEISS narrative. The analysis used a keyword-based definition for severity: any narrative containing terms indicative of admission/transfer or higher-acuity treatment (hospital, admit, ICU, inpatient, IV, IV fluids, transfer) was classified as severe. All other encounters were classified as not severe.

### Covariates and weights

We extracted patient age, sex, month and year of treatment, and NEISS-provided record weight. Year was centered for inclusion in models. Observations with missing Year were excluded. Frequency/sample weights supplied in the NEISS files were converted to numeric and used as frequency weights in regression models.

### Statistical analysis

We computed weighted descriptive statistics by GI status by summing NEISS weights and calculating weighted counts and weighted proportions of severe events. Yearly and seasonal rates were calculated as weighted severe events divided by weighted totals and expressed per 100,000 NEISS-weighted ED visits. Temporal trends were visualized with LOESS smoothing of yearly weighted rates and with heatmaps of monthly rates by year.

Association between GI infection and severe outcome was estimated using a weighted binomial generalized linear model (logit link). The primary model specification was:

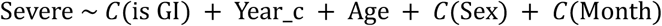

and was fit with statsmodels GLM (binomial family) using NEISS weights as frequency weights. Model-based (non-robust) standard errors and confidence intervals were reported from the fitted GLM object. All analyses and figures were produced with Python 3.10 (packages: pandas, NumPy, matplotlib, statsmodels).

### Analytic caveats

Because the analysis identified cases and severe outcomes using narrative keyword searches rather than diagnostic codes or laboratory confirmation, outcome and exposure misclassification are possible. In addition, although NEISS sampling weights were used as frequency weights in regression models, the analysis did not explicitly incorporate NEISS strata and primary sampling unit variables in a complex-survey variance estimator; consequently, the reported standard errors and confidence intervals are model-based and may under-or over-estimate true sampling variance.

## RESULTS

The analytic dataset included 2,876,152 pediatric emergency department encounters with non-empty narratives, representing an estimated 86.6 million weighted ED visits nationwide. Narrative keyword classification identified approximately 438,000 weighted visits as probable gastrointestinal (GI) infections. Severe outcomes—defined by narrative evidence of hospital admission, transfer, ICU-level care, or intravenous treatment—were uncommon overall but occurred disproportionately in GI-related encounters. Weighted estimates indicated 179,026 severe events among non-GI visits (0.21%) compared with 3,585 severe events among GI visits (0.82%), corresponding to roughly 817.9 severe events per 100,000 GI-related ED visits versus 206.8 per 100,000 non-GI visits (**Figure 1**). Temporal visualization showed relatively stable decline in early severe rates over the study period (**Figure 2**).

**Figure 1.**
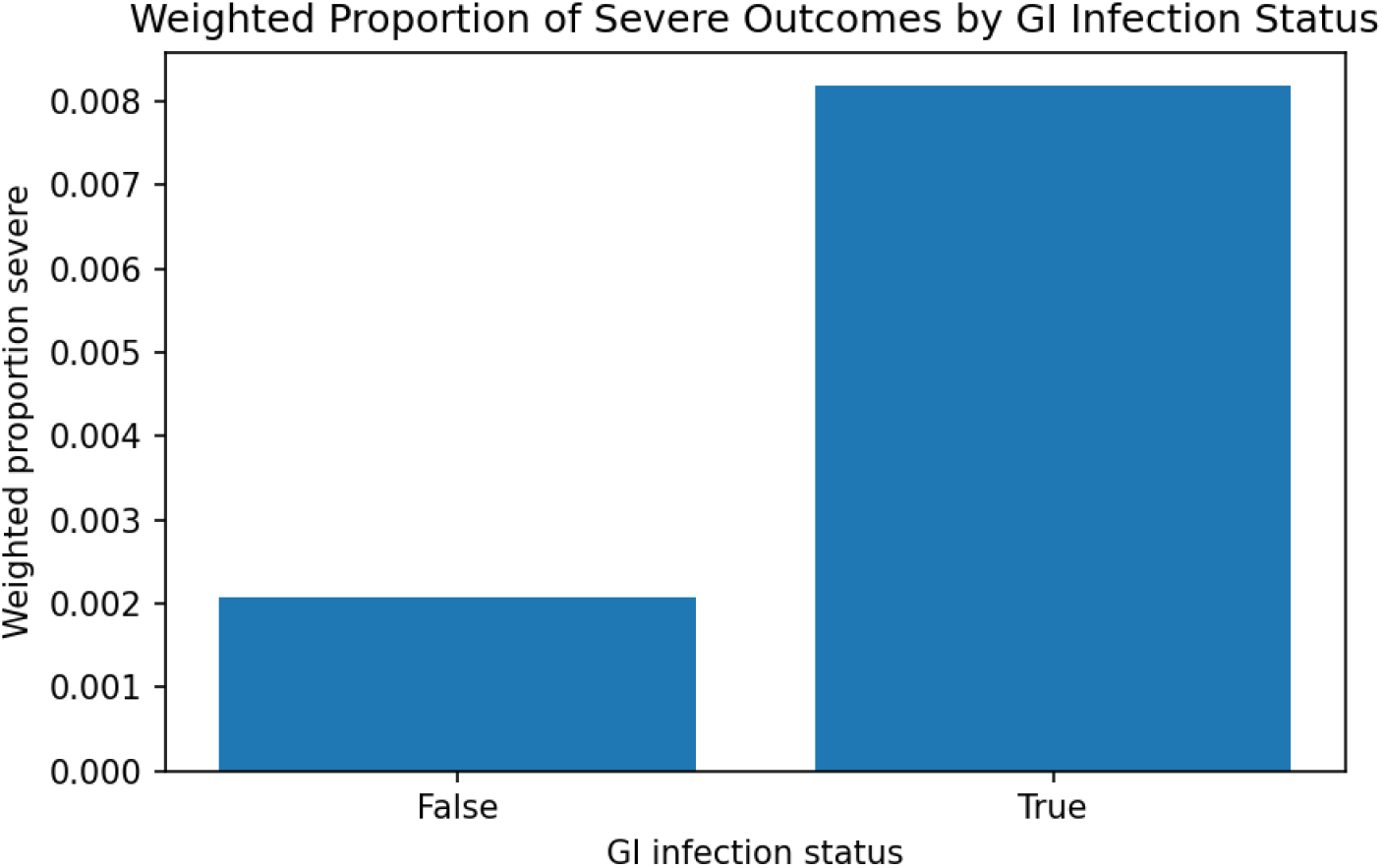
Weighted Proportion of Severe Outcomes by GI Infection Status.

**Figure 2.**
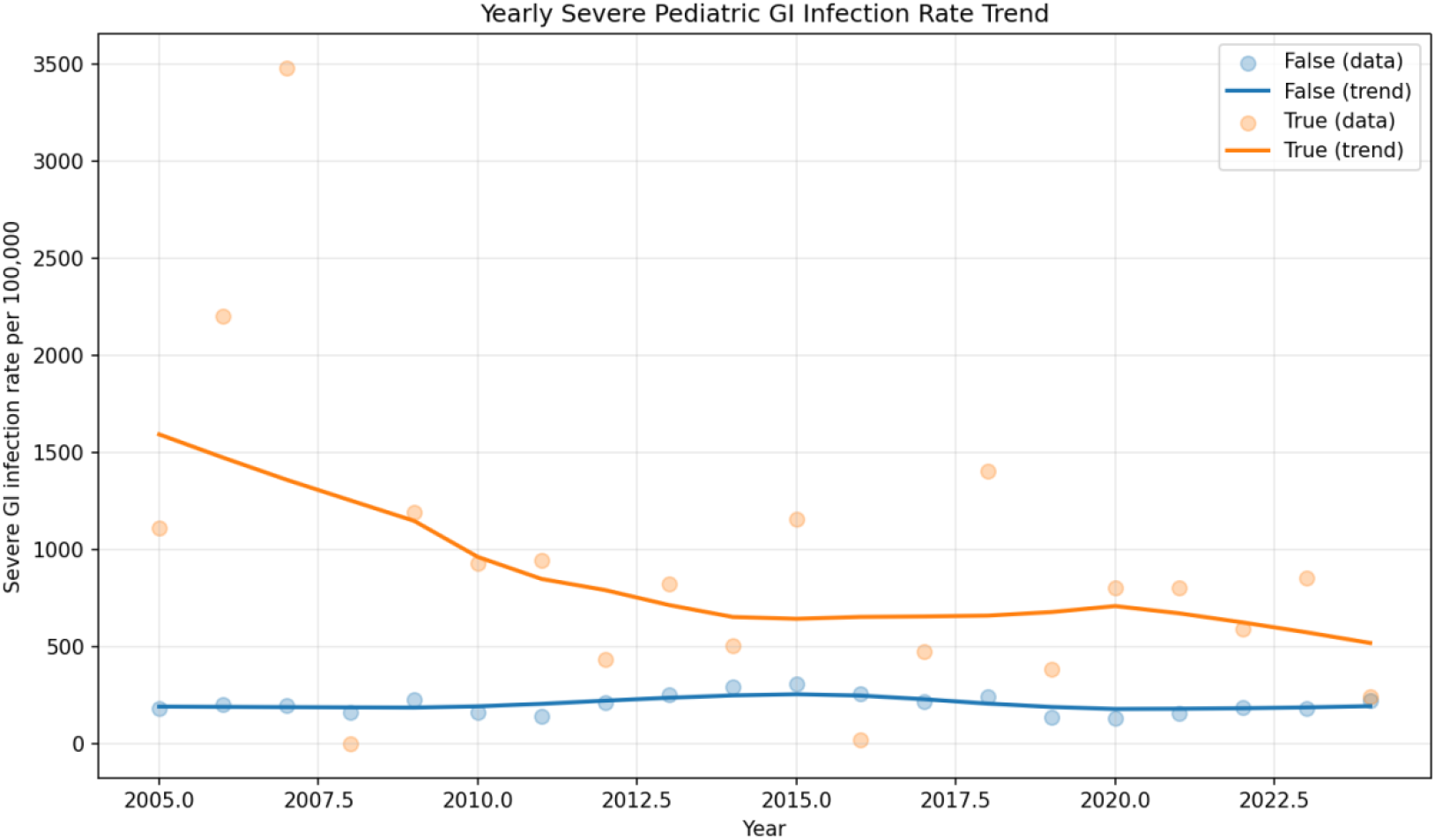
Yearly Severe Pediatric GI Infection Rate Trend.

In multivariable weighted regression adjusting for age, sex, calendar year, and month, GI infection status was strongly associated with increased odds of a severe outcome (adjusted OR 3.95, 95% CI 3.82–4.09, p < 0.001). Age was associated with slightly lower odds of severity (per-year OR ∼0.99, p < 0.001), and sex categories showed modest associations. Month effects aligned with seasonal patterns observed descriptively, with significantly higher odds in March and lower odds in May and December relative to the reference month. Calendar year did not demonstrate a significant linear trend after adjustment. Overall, GI-related pediatric ED visits exhibited a markedly higher burden of severe outcomes compared with other presentations, both in weighted proportions and in adjusted risk estimates.

## CONCLUSIONS

In this nationally weighted analysis of more than 2.8 million pediatric emergency department encounters—representing approximately 87 million ED visits nationwide— gastrointestinal infections emerged as a major driver of severe clinical outcomes in children. Despite their relatively low overall incidence, GI-related presentations accounted for disproportionately high rates of hospitalization, transfer, ICU-level care, or intravenous treatment, with nearly a fourfold increase in adjusted odds of severe outcomes compared with all other pediatric ED visits. This magnitude of risk, observed consistently across demographic groups and seasonal patterns, underscores that acute GI illness represents not merely a common complaint in emergency care, but a clinically consequential condition with meaningful implications for resource utilization and patient safety.

These findings elevate pediatric gastrointestinal infections as a significant and under-recognized contributor to severe emergency morbidity at the national level, particularly during seasonal surges in early spring when demand on acute care systems may already be strained. The identification of this elevated risk highlights the importance of rapid triage, early fluid resuscitation, and vigilant monitoring in children presenting with GI symptoms. While limitations inherent to narrative-based classification and NEISS surveillance data warrant continued refinement, the present study provides compelling national evidence that pediatric GI infections carry substantial clinical and public health impact. Strengthening surveillance, improving early recognition, and targeting prevention strategies have the potential to meaningfully reduce severe outcomes in this large and vulnerable patient population.

## Data Availability

All data produced in the present study are available upon reasonable request to the authors.

https://www.cpsc.gov/Research--Statistics/NEISS-Injury-Data

## DATA AVAILABILITY STATEMENT

All source data were openly available before the initiation of the study and were accessible at cpsc.gov/Research--Statistics/NEISS-Injury-Data. All data produced in the present study are available upon reasonable request to the authors.

## REFERENCES

1. Elliott EJ. Acute gastroenteritis in children. BMJ. Jan 6 2007;334(7583):35–40. doi:10.1136/bmj.39036.406169.80

2. Hasegawa K, Tsugawa Y, Cohen A, Camargo CA, Jr. Infectious Disease-related Emergency Department Visits Among Children in the US. Pediatr Infect Dis J. Jul 2015;34(7):681–5. doi:10.1097/INF.0000000000000704

3. Han SM, Duggan CP, Graham DA. Understanding the Burden of Pediatric Gastrointestinal Diseases-Does a Look From the Perspective of Inpatient Administrative Databases Help? J Pediatr. Mar 2018;194:11–12. doi:10.1016/j.jpeds.2017.11.034

4. Hamdan O, Stopczynski T, Amarin JZ, et al. 1748. Viral and Bacterial Etiology of Acute Gastroenteritis in Children < 5 Years Old at a Major Pediatric Referral Center Before and During the COVID-19 Pandemic. Open Forum Infectious Diseases. 2023;10(Supplement_2)doi:10.1093/ofid/ofad500.1579

5. Hallowell BD, Parashar UD, Curns A, Degroote NP, Tate JE. Trends in the Laboratory Detection of Rotavirus Before and After Implementation of Routine Rotavirus Vaccination — United States, 2000–2018. MMWR Morbidity and Mortality Weekly Report. 2019;68(24):539–543. doi:10.15585/mmwr.mm6824a2

6. Shah MP, Dahl RM, Parashar UD, Lopman BA. Annual changes in rotavirus hospitalization rates before and after rotavirus vaccine implementation in the United States. PLOS ONE. 2018;13(2):e0191429. doi:10.1371/journal.pone.0191429

7. Glass RI, Parashar U, Patel M, Tate J, Jiang B, Gentsch J. The control of rotavirus gastroenteritis in the United States. Trans Am Clin Climatol Assoc. 2012;123:36-52; discussion 53.

8. Burnett E, Parashar UD, Winn A, Tate JE. Trends in Rotavirus Laboratory Detections and Internet Search Volume Before and After Rotavirus Vaccine Introduction and in the Context of the Coronavirus Disease 2019 Pandemic—United States, 2000–2021. The Journal of Infectious Diseases. 2022;doi:10.1093/infdis/jiac062

9. Pines JM, Zocchi MS, Black BS, et al. Characterizing pediatric emergency department visits during the COVID-19 pandemic. Am J Emerg Med. Mar 2021;41:201–204. doi:10.1016/j.ajem.2020.11.037

10. Kim S, Ro YS, Ko SK, et al. The impact of COVID-19 on the patterns of emergency department visits among pediatric patients. Am J Emerg Med. Apr 2022;54:196–201. doi:10.1016/j.ajem.2022.02.009

